# Feasibility Study on Training Dogs to Detect Lung Cancer: Findings of a Retrospective Evaluation

**DOI:** 10.64898/2026.02.04.26345351

**Authors:** Christian Grah, Shiao Li Oei, Steven Ngandeu Schepanski, Hannah Wüstefeld, Katarzyna Blazejczyk, Julia Kalinka-Grafe, Georg Seifert

## Abstract

Early detection is critical for lung cancer patients. One lung cancer detection method under study is using sniffer dogs. This study aimed to evaluate, retrospectively, the sensitivity and specificity of the **C**ancer **D**etection **D**og **C**ollective (CDDC®) method under training conditions. A team of five trained sniffer dogs analyzed breath samples from lung cancer patients and cancer-free volunteers, and a cancer sample is positive if at least three dogs indicate it. Dog handlers and experimental observers were blinded to sample identity, and detection accuracy was assessed. Primary endpoint was sensitivity, and selectivity and confounding factors were also assessed. Samples were collected in 2024 from 824 volunteers, including 111 with a confirmed diagnosis of lung cancer (mean age 60, range 34-80, 18% early-stage cancer, 46% not yet oncological treated). A total of 11,900 breath samples were tested with 125 test runs per dog. Individually, the five dogs demonstrated detection performance with sensitivities between 82% and 89%, and specificities of over 95%. The CDDC® dog team’s corporate decision revealed a sensitivity over 95% and the rate of false positives was 0%. Analysis of potential confounding factors revealed that weather conditions and supervisor skills were associated with the dogs’ performance. The CDDC® method showed high consistency in training scenarios. Further studies should evaluate this method in a controlled clinical study alongside lung cancer screening.

## 1. Introduction

Lung cancer is still the leading cause of cancer death worldwide. Late detection contributes to poor treatment outcomes, and the early detection of lung cancer continues to be a challenge [1]. Lifestyle changes, such as smoking cessation, have been shown to significantly reduce cancer-specific mortality in lung cancer survivors [2]. For smokers, quitting remains the most effective way to prevent cancer and cardiovascular disease [3]. Reducing the mortality rate for lung cancer may be possible through early detection using low-dose computed tomography (LDCT) in high-risk individuals. Many countries are making progress in establishing national screening programs [4]. However, lung cancer screening with LDCT can result in false positive test results, leading to stressful examinations for patients [5]. Another innovative approach involves training animals to detect cancer, a method that has demonstrated promising results [6]. The use of the dog’s sense of smell is well known, researched, and published [7-9]. Published studies have reported a sensitivity of 78% and a specificity of 71.5% for the detection of lung cancer by sniffer dogs in breath and urine samples [10-13]. The exact chemical compounds or combinations of compounds that dogs sniff that indicate lung cancer have not been determined. It has also not yet been confirmed whether dogs can detect early, preclinical stages of cancer with comparable accuracy to already diagnosed cases [9]. A total of 62 studies were analyzed in a meta-analysis review [14], but the available studies showed a very heterogeneous picture with regard to study quality. The risk of bias in the studies was often high, resulting in low-quality evidence. In a double-blind clinical study using breath and urine samples, the dogs identified 40 of 41 lung cancer samples from a total of 246 patients, corresponding to an overall detection rate of 97.6% [15]. Another study found that using exhaled breath for training sniffer dogs is more sensitive and specific than using urine or tissue samples [16]. This training method had a 91.7% sensitivity and an 85.1% specificity [16]. In another review, it was concluded that medical scent canines could be comparable in sensitivity to LDCT lung cancer screening, and may even have higher specificity [17]. Further refinement of this research could potentially lead to cancer screening tests being carried out in the future.

In present evaluation, a newly refined Cancer Detection Dog Collective (CDDC®) method was evaluated, with the aim of optimizing breath sampling for ease of use and reproducibility. To increase the detection rate, the results of previously published methods [10,11] were considered and optimized so that the CDDC® method includes five independent test runs with five different dogs.

The aim of this retrospective evaluation was to determine the sensitivity and specificity of the CDDC® test under training conditions.

## 2. Materials and Methods

### 2.1. Study design

This was a retrospective evaluation of a dog-based cancer detection procedure. Positive breath samples from lung cancer patients and negative breath samples from presumably healthy persons were used for all the training runs. Lung cancer patients were recruited from various German hospitals and written informed consent was obtained from all subjects, who provided clinical lung cancer diagnoses and breath samples for research purposes. No compensation was given to the subjects who provided breath samples. All animal-training and -handling methods were designed in consultation with veterinarians, dog trainers, and dog owners. All procedures were carried out by Dogscan GmbH in Erkelenz, Germany (dogscan-deutschland.de). Ethical approval for the retrospective evaluation and analysis of the anonymized data was obtained from the ethics committee of Charité – Universitätsmedizin Berlin (EA1/111/25) on 22.05.2025.

### 2.2. Sample collection

Between January and September 2024, breath samples of 824 volunteers were collected at multiple locations in Germany. The breath samples from lung cancer patients were clinically confirmed and provided by lung cancer centers and hospitals, including Luisenhospital Aachen and Elisabeth-Hospital of the Städtische Kliniken Mönchengladbach, as well as other oncology or pulmonology specialist practices in Germany. Volunteers without lung cancer were recruited from among students, as well as from people with preexisting respiratory diseases for whom lung cancer had been ruled out by clinical examination. All participants signed a consent form agreeing to provide breath samples and make their data available. The use of fleece masks for sampling exhaled volatile organic compounds has been recommended for canine scent tests [18]. Here, we used certified (Article 2, No. 3 of Regulation (EU) 2017/746 (IVDR)) breathing masks (100% cotton, OEKO-TEX® Standard 100, Product Class I (Baby Class), with integrated absorbers (100% cotton) for aerosols. Each participant was instructed to wear the mask for five consecutive minutes, breathing out exclusively through their mouth. All breath samples were collected outdoors, away from clinical rooms and in an environment with extraneous odors. Thereafter, the masks were immediately sealed in airtight foil bag and stored at ambient temperature (10°C-30°C). The masks were analyzed within a maximum of three weeks after they were worn.

### 2.3. Dog training, testing and detection procedure

Family dogs were trained by professional dog trainers with experience in scent detection. Sniffer dog training protocols were adapted from other studies [10] and optimized for the present setting. Similar to what is described by others [10, 15-17], young puppies were trained in several stages to recognize and indicate breath samples from lung cancer patients. From the start of their training, the dogs are trained and tested using control masks with strong, distracting odors, such as food, smoke, or cosmetics, to ensure they can detect the target odor specifically. Positive breath samples from lung cancer patients with clinically confirmed diagnoses (various stages, histologies, and molecular pathologies) were also used for the training and education of the sniffer dogs. Methodological standards for olfactory detection trials were followed as recommended [9] and the performance of the dogs was verified by a rigorous double-blind (dog and dog handler) procedure. All of the sniffer dogs used here undergo extensive training, must have at least 12 months of experience in detecting lung cancer breath samples. The sniffer dogs must regularly demonstrate their detection performance and sniff out a cumulative total of 2,500 samples, including 100 breath samples from individuals with a confirmed lung cancer diagnosis. The detection performance is monitored by an independent body, and the minimum requirements are sensitivity of 90% and specificity of 99.13%.

In this study, five trained sniffer dogs were used see Figure 1, three of which were male and two females, with ages ranging from two to seven years. Dog training was performed in a specialized facility equipped with standardized search boards as shown in Figure 1. Unlike previous test procedures conducted under controlled laboratory conditions, sterile search containers were not used here. Instead, numerous search boards, up to 200 were set up in a natural environment with foreign odors. The number of search boards and arrangement of samples was determined by a computer-assisted randomization algorithm (Ninox Enterprise, Customized Programming, Germany). The supervisor, who was independent of the experimental observers and dog handlers, was responsible for preparing the search layout and placing the samples. All observers and dog handlers were blinded, and the supervisor was the only person who knew where the positive and negative masks were placed. The supervisor remained at a constant distance of 8–12 meters from the search boards during the search runs, standing in the same position to avoid position-related stimuli. If the searching dog indicates a mask, the respective dog handler says “indication,” and if it is a positive mask the supervisor answers “*Proband*” and the dog receives a reward. If the dog indicates a negative mask, the supervisor answers “*misleading*” and the dog handler leads the dog on without giving a reward. The supervisor documents all displayed search board numbers in the system log. Dog handlers were unaware of sample placement and the independent observers were blinded to sample status throughout the trial. For each search run, 25 to 200 search boards equipped with different breathing masks are set up in the test facility as shown in Figure 1.

**Figure 1.**
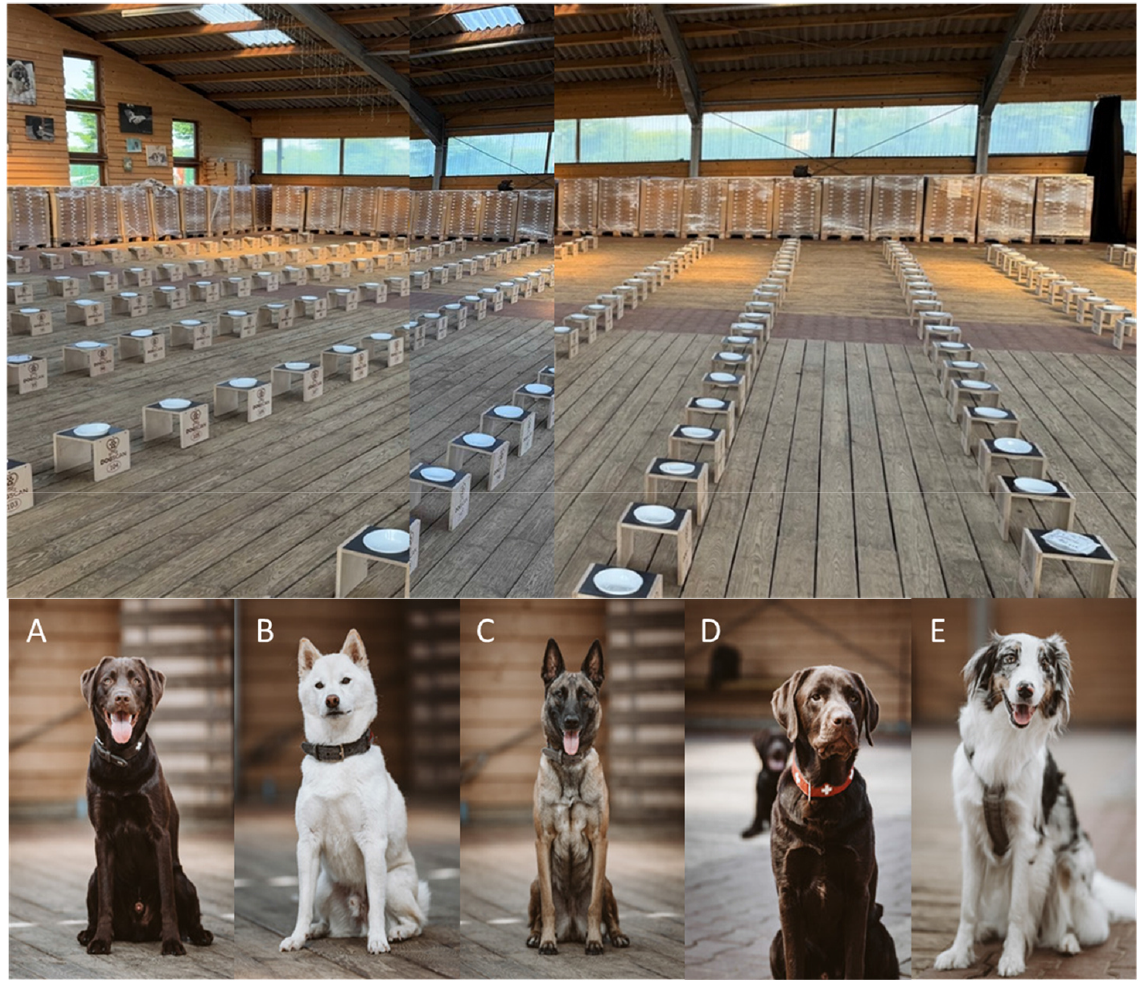
The Cancer Detection Dog Collective Team. The test facility with search boards is shown above. Below are the five sniffer dogs used: **A)** male Labrador, 2 years, **B)** male, Shiba Inu, 7 years, C) female, Belge Malinois, 4 years, **D)** male, Labrador 3 years, **E)** female Australian Shepherd, 3 years.

A typical search run consists of 100 search boards, five of which are populated with lung cancer breath samples (P) and 95 of which are populated with masks from non-cancer subjects (N). A positive indication of lung cancer was defined as the dog stopping in front of a sample, while a negative indication was defined as the dog running past a sample. A typical test run with 100 search boards, including five positive samples, takes about eight minutes per dog. At least two independent observers recorded and scored each dog’s indications and documented the numbers of true positive (TP), true negative (TN), false positive (FP), and false negative (FN) per run. All sniffer dogs undergo regular training, with ten to fifteen test runs each month. Fresh samples are used in these searches, and the dogs’ performance is evaluated and documented.

### 2.4. Statistical analysis

All statistical analyses were conducted using Python (version 3.11.3) with the following libraries: pandas, numpy, scikit-learn, statsmodels, and matplotlib. A significance threshold of p < 0.05 was applied throughout the analysis. Participant characteristics and sample distributions were summarized using descriptive statistics.

For each test run and dog, we counted true positives (TP), false positives (FP), true negatives (TN), and false negatives (FN) responses and performance metrics were estimated. Sensitivity is calculated as TP / P and specificity as TN / N. Exact 95% confidence intervals (CIs) for detection metrics were estimated using the Wilson score method, and corresponding confusion matrices and classification distributions were generated. Since repetitive samples from the same donors were used, corrected metrics were calculated to account for potential pseudoreplication bias. The number of false negative (FN) and false positive (FP) values were counted as before. The corrected s*ensitivity* and *specificity* were recalculated using the number of unique subjects. *P* = unique number of P; *N* = unique number of N; *Sensitivity* = (*P* - FN)/*P*; *Specificity* = (*N* - FP)/*N*.

In order to assess the robustness of the ensemble decision process, the number of dogs involved was varied. To this end, a stratified bootstrapping procedure was implemented, based on a mask-by-dog detection outcome matrix. For each scenario (e.g., using 2, 3, or 4 dogs), we randomly sampled the specified number of dogs without replacement and applied a modified collective decision rule (e.g., mask is positive if ≥ 1 of 2, ≥ 2 of 3, or ≥ 3 of 4 dogs indicate it). This process was repeated 5,000 times per scenario to obtain stable performance estimates. Across iterations, we computed the distribution of sensitivity, specificity, and accuracy metrics, as well as their standard deviations (SD), to assess their reliability of varied configurations.

For evaluating confounding variables, we used a generalized estimating equation (GEE) logistic regression model with a binomial family to account for clustering by dogs. Evaluated variables included ambient temperature, weather conditions, dog mood (as rated by handlers), and supervisor identity. Robust standard errors and regression coefficients with 95% CIs were computed.

## 3. Results

Between January and September 2024 breath samples from a total of 824 subjects, including 111 individuals with a histologically confirmed lung cancer and 713 volunteers with no prior lung cancer history were included in the evaluation. The characteristics of the donors of breath samples included in the study are presented in Table 1.

**Table 1.** Characteristics of the patient collective.

| n= 111 (100%) |  | Lung cancer |  | n= 713 (100%) |  | Control |
| --- | --- | --- | --- | --- | --- | --- |
| age in years, mean, range |  | 60 (34-80) |  | age 18-23, n (%) |  | 526 (74%) |
| gender, male, n (%) |  | 23 (21%) |  | age 24-28, n (%) |  | 98 (14%) |
| female, n (%) |  | 51 (46%) |  | age 29-65, n (%) |  | 61 (9%) |
| unknown, n (%) |  | 37 (33%) |  | age 65+, n (%) |  | 28 (4%) |
| ever smoker, n (%) |  | 12 (11%) |  | ever smoker, n (%) |  | 213 (30%) |
| never smoker, n (%) |  | 13 (12%) |  | never smoker, n (%) |  | 342 (48%) |
| unknown, n (%) |  | 87 (78%) |  | unknown, n (%) |  | 158 (22%) |
| **Histology, n (%)** |  |  |  | **Student Cohort** |  | 528 (74%) |
| non-small cell lung carcinoma (NSCLC) |  | 89 (80%) |  | **Chronic Disease Cohort** |  | 185 (26%) |
| - adenocarcinoma |  | 58 (52%) |  | - respiratory diseases (asthma, COPD, pulmonary emphysema) |  | 59 (8%) |
| - squamous cell carcinoma |  | 14 (13%) |  | - other chronic diseases |  | 126 (18%) |
| - other or unknown |  | 17 (15%) |  |  |  |  |
| small cell lung cancer (SCLC) |  | 7 (6%) |  |  |  |  |
| other or unknown histology |  | 15 (14%) |  |  |  |  |
| UICC stage: stage I, n (%) |  | 10 (9%) |  |  |  |  |
| stage II, n (%) |  | 8 (7%) |  |  |  |  |
| stage III, n (%) |  | 16 (14%) |  |  |  |  |
| stage IV, n (%) |  | 42 (38%) |  |  |  |  |
| brain metastases, n (%) |  | 3 (3%) |  |  |  |  |
| unknown, n (%) |  | 35 (32%) |  |  |  |  |
| **Received treatment: n, (%)** |  |  |  |  |  |  |
| untreated |  | 51 (46%) |  |  |  |  |
| radiation |  | 21 (19%) |  |  |  |  |
| chemotherapy |  | 45 (41%) |  |  |  |  |
| immunotherapy |  | 29 (26%) |  |  |  |  |

**Abbreviation:** COPD: Chronic Obstructive Pulmonary Disease; UICC: Union for International Cancer Control

Ten percent of the lung cancer patients recruited were classified as Union for International Cancer Control (UICC) stage I, and 8% were classified as stage II. This indicates that 18% of the patients were in the early stages of lung cancer. Forty-six percent of the lung cancer patients had not undergone oncological treatment prior to donating positive breath samples (Table 1). The majority of volunteers who provided negative breath samples were between 18 and 23 years old, in good health, and free of known diseases. The 713 control subjects can be grouped into two cohorts: The first is the “student cohort,” which consists of 528 healthy control subjects who are mostly young. The second is the “chronic disease cohort” (26%), which consists of 185 non-cancer patients with confirmed chronic diseases (See Table 1). 59 patients of the chronic disease cohort suffered from chronic respiratory diseases and in further 126 cases from other chronic diseases but not from cancer (Table 1).

### 3.1. Dogs detection performance

Each dog conducted 125 search runs with in total 592 positive and 11,308 negative samples. Figure 2 shows an overview diagram of the number of included breath masks, test boards, and dog evaluations. A total of 592 positive (P) and 11,308 negative (N) masks were tested per dog in parallel. A total of 125 test runs per dog were carried out within nine months. The tests were carried out by five dogs A-E in Figure 1, resulting in a total of 625 test runs and 59,500 mask evaluations as shown in Figure 2.

**Figure 2.**
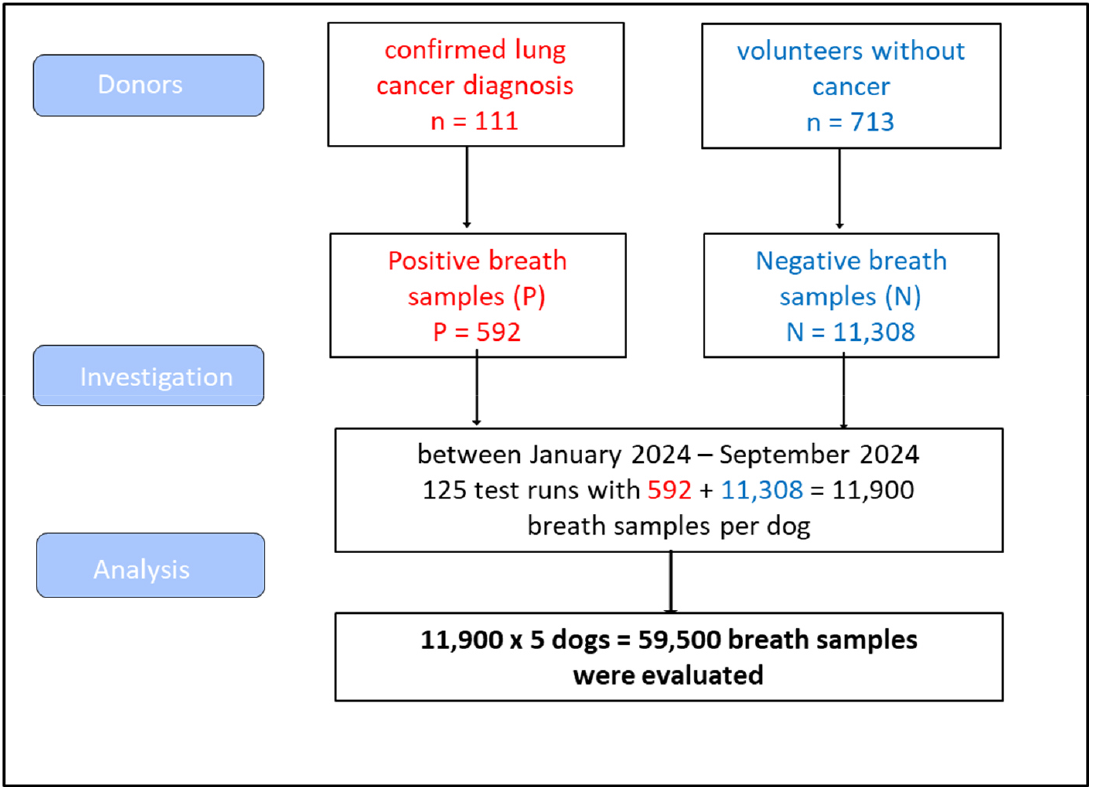
Overview of included breath samples and dog evaluations.

The performance metrics were calculated separately for each dog based on standard diagnostic categories with 95% CI using the Wilson score method. A total of 625 independent test runs were performed and in summary the following results were obtained (Table 2). All five dogs demonstrated comparable detection performance, sniffing out a total of 11,900 breath samples each, with a sensitivity of above 96% and a specificity of above 99%. The recalculated performance metrics for the five dogs, based on the number of unique subjects (*P* = 111; *N* = 713), resulted in *sensitivities* ranging from 82% to 89% and *specificities* greater than 95% (Table 2). The resulting false positive rate was below 5% per dog.

**Table 2.**
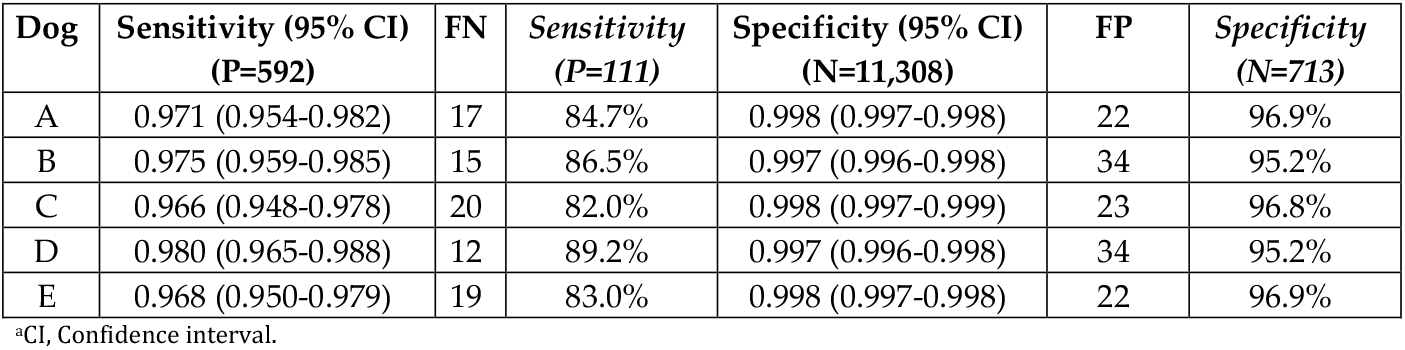
Dog’s individual detection performance.

### 3.2. Dogs detection performance

A bootstrap resampling analysis was performed with 10,000 iterations per configuration. For each iteration, subsets of two, three, or four dogs were randomly selected from the 625 test runs (see Table 3). All simulated dog team sizes demonstrated robust sensitivities (Table 3).

**Table 3.** Detection performance for different dog team sizes.

| Dog Team Setup | Sensitivity (95% CI<sup>a</sup>)<br>(P=592) | FN | Sensitivity<br>(P=111) |
| --- | --- | --- | --- |
| 2 dogs (cutoff ≥1) | 0.994 (0.990-0.997) | 4 | 96.4% |
| 3 dogs (cutoff ≥2) | 0.990 (0.986-0.995) | 6 | 94.6% |
| 4 dogs (cutoff ≥3) | 0.985 (0.981-0.990) | 9 | 91.9% |
| 5 dogs (cutoff ≥3) | 0.992 (0.992-0.992) | 5 | 95.5% |

Figure 3 provides a summary of the combined results according to the CDDC® rules. The evaluation of all 125 independent test runs were calculated using the number of unique donors, 111 positive *(P)* and 713 negatives *(N)*. The results according to the CDDC® rules were zero false positive (FP), two false negatives (FN), and 131 inconclusive results (one or two dogs indicated the sample). Accordingly, the five dogs CDDC® test achieved at least a *sensitivity* of 95.5%. In 128 cases one or two of the five dogs indicated a control sample as FP, thus the *specificity* for that control samples were recognized as true negatives by all five dogs was at least 82%. According to CDDC® rules, none of the 11,308 negative samples were classified as FP, implying that the false positive rate is 0%.

**Figure 3.**
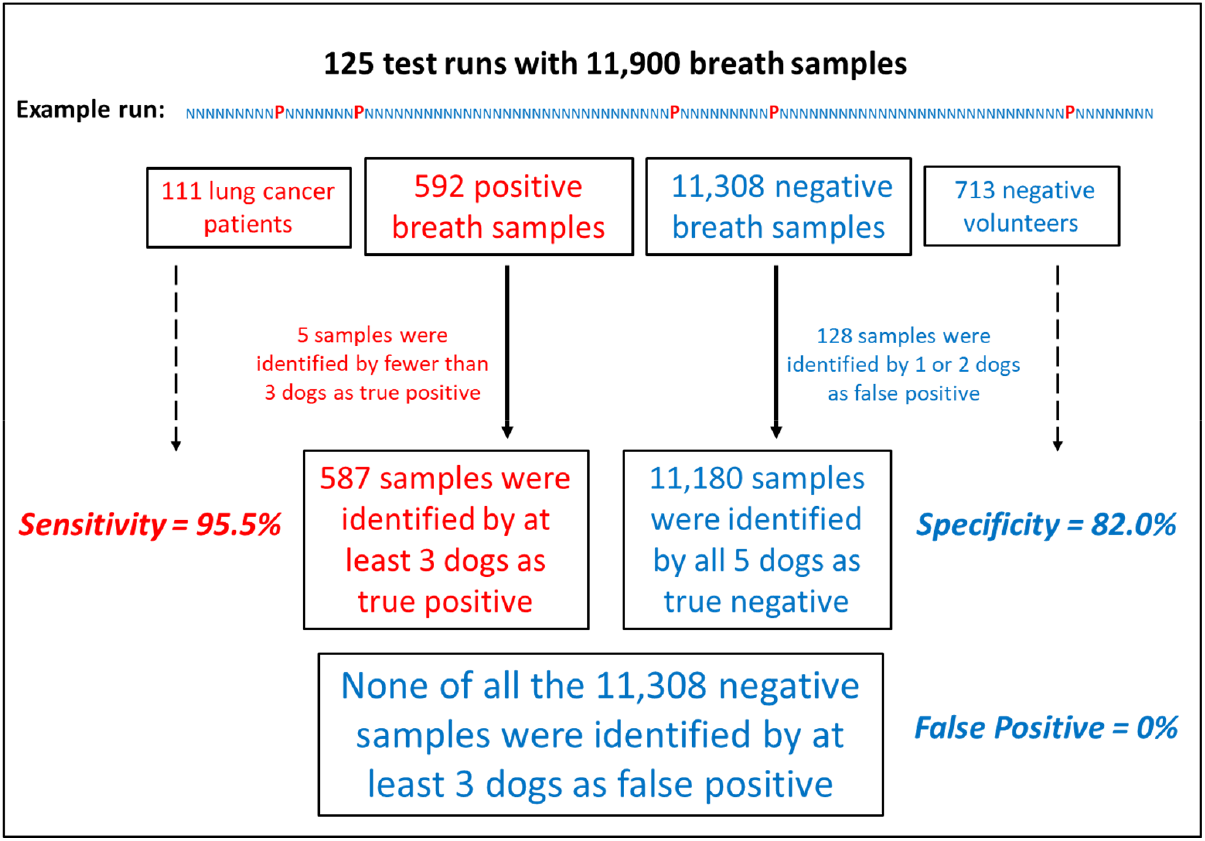
Summary of the five dogs’ evaluation according to the CDDC® rules

### 3.3. Model assumption for preexisting comorbidities (explorative sensitivity analysis)

Most negative donors were young and healthy (the student cohort), but 26% of the control subjects had pre-existing chronic diseases (see Table 1). This group with chronic diseases is a more suitable comparison group; a sub analysis was performed, limiting the model assumption to the Chronic Disease Cohort. For this purpose, all counted false positives (FP) were assigned to the subjects of the Chronic Disease Cohort (N = 185). Thus, the *specificities* for the dogs’ individual detection performance, as displayed in Table 2, were calculated as (185 – FP)/185. This would result in *specificities* ranging from 81.6% to 88% for the five dogs. With regard to the CDDC®-Rules (compare Figure 3), the resulting False Positive rate for the Chronic Disease Cohort still would remain at 0%.

### 3.4. Analysis for potential confounders on dogs’s detection performance

To evaluate potential predictors and confounding influences on detection performance, we fitted generalized estimating equations models (Table 4). The model incorporated dog mood, weather conditions, ambient temperature, and the identity of the supervising person as independent variables. The analysis was based on each dog’s detection performance per trial day, yielding a total of 625 dog-day observations across 125 trial days.

**Table 4.** Association factors for detection performance.

| 125 search runs x 5 dogs- n=625 (100%) |  |  |  |
| --- | --- | --- | --- |
| Predictor | Coefficient ($\beta$) | 95% CI<sup>s</sup> | p-value |
| **dog mood** | Reference: balanced mood, n=538 (86%) |  |  |
| Motivated, n=52 (8%) | 0.776 | (-0.169, 1.722) | 0.108 |
| overly motivated, n=35 (6%) | -0.203 | (-0.897, 0.491) | 0.566 |
| **weather** | Reference: sunny, n=185 (30%) |  |  |
| Cloudy, n=100 (16%) | -0.947 | (-1.490, -0.403) | 0.001* |
| Rainy, n=340 (54%) | -0.008 | (-0.595, 0.578) | 0.978 |
| **temperature**<br>range 4°C–32°C; mean 15.4°C | 0.0004 | (-0.031, 0.031) | 0.981 |
| **supervisor** | Reference: person 1; n=275 (44%) |  |  |
| Person 2, n=45 (7%) | -0.324 | (-0.597, -0.052) | 0.020* |
| Person 3, n=85 (14%) | -0.071 | (-0.582, 0.439) | 0.439 |
| Person 4, n=100 (16%) | -0.223 | (-0.599, 0.153) | 0.245 |
| Person 5, n=10 (2%) | -1.345 | (-2.409, -0.282) | 0.013* |
| Person 6, n=35 (6%) | 0.966 | (0.088, 1.843) | 0.031* |
| Person 7, n=20 (3%) | 0.425 | (-0.967, 1.816) | 0.550 |
| Person 8, n=15 (2%) | -0.596 | (-0.951, -0.240) | 0.001* |
| Person 9, n=20 (3%) | 0.025 | (-1.02, 1.07) | 0.962 |
| Person 10, n=20 (3%) | 1.570 | (-0.529, 3.669) | 0.143 |
| <sup>s</sup>CI, Confidence interval; n, number of search runs. Negative coefficient $\beta$ indicate an association with lower detection performance, while positive coefficient $\beta$ indicate an association with better performance. *Significant p-values $p < 0.05$ are indicated. | | | |

Under reference conditions, balanced dog mood, sunny weather, and average ambient temperature of 15.35 °C, and supervisor person 1, the model intercept was significantly positive (β = 1.291, p < .001), reflecting high baseline detection accuracy. The dog’s mood, as reported by the handler, and the ambient temperature do not appear to have a significant impact on the dogs’ detection performance, while cloudy in relation to sunny weather appears to significantly reduce detection performance (β = –0.95, p = .001). A discernible variation in the detection performance of the dogs was observed regarding the different supervisors of the different search runs. Person 2 who supervised 7% of the search runs, was associated with reduced detection performance (β = -0.324, p = .020). Similarly, persons 5 and 8, who each supervised only 2% of the trials, were significantly negatively associated with detection performance (β = -1.345, p = .013 and β = - 0.596, p = .001, respectively). In contrary, person 6 was associated with improved performance (β = 0.966, p = .031). No statistically significant differences in detection performance were found among the other supervisors.

## 4. Discussion

Under training conditions, detecting lung cancer from breath samples using the CDDC® method with five dogs appears feasible, with at least 95.5% sensitivity and no false positives. The minimum requirement for trained sniffer dogs is a sensitivity of at least 90%. Previous and more recent studies involving sniffer dogs have shown that dogs are generally capable of detecting lung cancer breath samples with high sensitivity and specificity [10, 12, 15-17, 19]. Ehmann et al. (2012) proposed using a “corporate decision” strategy with multiple dogs to improve sensitivity since hit rates vary among individual dogs. We were indeed able to demonstrate that the CDDC® method increases sensitivity. Additionally, the CDDC® rule ensures that a sample is only considered negative if none of the five dogs react to it, ensuring that false positive and false negative rates remain very low. However, since dogs are living beings and not machines, maintaining this level of sensitivity and specificity is difficult over long periods of time. This requires thoroughly thought-out and carefully structured training of the dogs. Notably, in present evaluation, the analysis of potential confounding factors revealed that neither the dogs’ mood nor temperature had a significant influence on the dogs’ detection performance. Despite belonging to different breeds (see Figure 1), the dogs showed no significant differences in sensitivity or selectivity in their detection performance (Table 2). However, a significant association between cloudy weather and reduced performance was identified (Table 4). Additionally, significant associations were identified between the dogs’ detection performance and the individual supervisors responsible for coordinating the search runs (see Table 4). The supervisor’s tasks are complex and require experience and routine to achieve a reliably high sensitivity value. For instance, Persons 2, 5, and 8, which coordinated only 2-7% of the search runs, were associated with significantly lower detection performance. On the other hand, with Person 6 in charge of supervision, this was seemingly associated with best detection performance (see Table 4). The dogs’ detection performance partly depends on the supervisors’ communication skills, so the search runs’ optimization is possible. Interestingly, another study [19] recently demonstrated the high-performance skills of sniffer dogs in detecting cancer samples. Unlike in our study, they used samples of different types of cancer and analyzed the probes by combining trained detection canines with artificial intelligence (AI) tools. Dogs were allowed to identify six samples one by one in a special detection room. The dogs’ reactions were monitored and analyzed with the assistance of AI. Notably, their results showed that incorporating AI increased sensitivity from 64.4% to 94.8%. The analyses of potential confounders performed here suggest that the dog search runs should be directed by an experienced supervisor, and should be avoided in cloudy weather.

The CDDC® method demonstrated high consistency in the training scenarios evaluated here. In the context of planned future screenings using authentic samples of suspected cancer, further improvements and quality control are required. When training sniffer dogs, it is important to avoid establishing recognizable patterns. The dogs should not be conditioned to recognize typical hospital odors. Therefore, all breath samples, including those from lung cancer patients, must be collected in a neutral environment, such as outdoors. Training and education of the dogs will also be further improved. The dogs’ well-being will be prioritized in the future development of this method, ensuring high-accuracy detection in natural, stress-free conditions. To ensure quality control, process optimization with regular monitoring is planned. Temperature, humidity, and air pressure affect the volatility, release, and dispersion of odors. Approaching rain can also activate the release of aromas from the environment, which can interfere with accurate detection. In order to limit the dependence of the accuracy of search runs on weather conditions, constructive measures should be taken at the facility. In addition to early detection, smoking cessation is an important way to reduce the risk of cancer and is a mandatory component of lung cancer screening programs [20] and should not be overlooked in the future.

This study has numerous limitations. One major limitation is its retrospective study design, as well as the fact that the control group was not age-matched. Also, complete demographic data and smoking status information was unavailable for all subjects. Furthermore, only breath samples from patients with confirmed lung cancer diagnoses were used for training and unclear suspicious lung cancer cases were not tested. However, a recalculation of all performance metrics based on the number of unique subjects, as well as an exploratory sensitivity analysis, showed high detection accuracy even when considering only subjects with chronic respiratory diseases like asthma or COPD, but not cancer.

It is still unclear whether the dogs can detect suspected cases or early-stage lung cancer within a lung cancer screening program. Furthermore, the use of sniffer dogs is limited by their inherent nature. Because they are living beings, not machines, the performance of sniffer dogs can be influenced by a wide variety of factors. For this reason, doubts have been raised about their suitability for reliable use in clinical practice [14]. Many of these uncertainties were here identified, managed, and had been optimized during the rigorous development of the CDDC® method’s training and implementation protocol. Even if this dog scent method may not be suitable for the direct early detection of lung cancer as part of a screening program, it has the potential to reduce the number of false-positive CT findings and to confirm a lung cancer diagnosis. Furthermore, it could support additional examinations and monitoring during the course of anticancer treatment. Additionally, the use of CT scans, which consume a lot of energy, as well as patient exposure to radiation, could potentially be reduced.

## 5. Conclusion

The reduction in lung cancer mortality is be achievable through screening of high-risk individuals and accordingly national screening programs with LDCT are in progress in many countries [4]. The CDDC® method exhibits several noteworthy advantages when considered as a potential cancer screening method. It is non-invasive, has no adverse effects, the breath sampling, storage, testing is relatively simple, and the evaluation of the results is straightforward. Labradors, Retrievers, and German Shepherds seem to be particularly good at scent detection [8], so these dogs have been trained here. This method differs considerably from other canine detection methods. First, living with dogs as family members means that dog and dog owner always work together as a unit. Second, they work in a “naturally contaminated” environment with foreign odors instead of clean sterile rooms. Third, they examine 100 samples in a single scan instead of just few samples in sterile boxes. Fourth, each search run is guaranteed to have quality assurance with accompanying positive and negative control masks. It has been proposed that a suitable screening method must have a true negative rate close to 100% to be considered reliable for safe use [21]. The sensitivity achieved here of at least 95.5%, coupled with the absence of false positive results in the corporate decision of the dogs, appears to meet these requirements. However, this present retrospective evaluation only used breath samples that were clearly positive or negative and only a minority of suitable control subjects without lung cancer were included. Therefore, it is still unclear whether dogs can detect suspected or early-stage lung cancer cases as part of a screening program. Thus, to validate the detection accuracy of the CDDC® method for the early detection of lung cancer, it is essential to prospectively investigate true, clinically conspicuous cases of suspected lung cancer in a controlled clinical study with direct comparison to parallel CT examinations. Considering the guidelines for the detection of lung cancer [22] and the recently published White Paper on Assessment of Potential Participants in Lung Cancer Screening in Germany [23], the CDDC® method will be tested prospectively in a planned proof-of-principle study. Recruiting suitable non-cancer subjects will be a major challenge for the future study.

## Acknowledgements

We would like to thank the whole team at Docscan GmbH in Erkelenz Germany, the dog trainers, and the dog owners for their excellent support in providing data. Special thanks go to the scent dogs, Aaron, Aki, Biest, Loki, and Lu. We would also like to thank all the volunteers who supported this study by donating their breath samples.

## Institutional Review Board Statement

The study was conducted in accordance with the Declaration of Helsinki, and the retrospective evaluation of this analysis was approved by the ethics committee of Charité – Universitätsmedizin Berlin (EA1/111/25) on 22.05.2025.

## Data Availability Statement

All data generated or analysed (https://github.com/sschepanski/CanineLungCancerDetection) during this study are included in this article.

